# Localising Vaccination Services: Qualitative Insights on an Orthodox Jewish Collaboration with Public health during the UK coronavirus Vaccine Programme

**DOI:** 10.1101/2021.09.10.21263372

**Authors:** Ben Kasstan, Sandra Mounier-Jack, Louise Letley, Katherine M Gaskell, Chrissy H Roberts, Neil RH Stone, Sham Lal, Rosalind M Eggo, Michael Marks, Tracey Chantler

## Abstract

Ethnic and religious minorities have been disproportionately affected by the SARS-CoV-2 pandemic and are less likely to accept coronavirus vaccinations. Orthodox (Haredi) Jewish neighbourhoods in England experienced high incidences of SARS-CoV-2 in 2020-21 and measles outbreaks (2018-19) due to suboptimal childhood vaccination coverage. The objective of our study was to explore how the coronavirus vaccination programme (CVP) was co-delivered between public health services and an Orthodox Jewish health organisation.

Methods included 28 semi-structured interviews conducted virtually with public health professionals, community welfare and religious representatives, and household members. We examined CVP delivery from the perspectives of those involved in organising services and vaccine beneficiaries. Interview data was contextualised within debates of the CVP in Orthodox (Haredi) Jewish print and social media. Thematic analysis generated five considerations: i) Prior immunisation-related collaboration with public health services carved a role for Jewish health organisations to host and promote coronavirus vaccination sessions, distribute appointments, and administer vaccines ii) Public health services maintained responsibility for training, logistics, and maintaining vaccination records; iii) The localised approach to service delivery promoted vaccination in a minority with historically suboptimal levels of coverage; iv) Co-delivery promoted trust in the CVP, though a minority of participants maintained concerns around safety; v) Provision of CVP information and stakeholders’ response to situated (context-specific) challenges and concerns.

Drawing on this example of CVP co-delivery, we propose that a localised approach to delivering immunisation programmes could address service provision gaps in ways that involve trusted community organisations. Localisation of vaccination services can include communication or implementation strategies, but both approaches involve consideration of investment, engagement and coordination, which are not cost-neutral. Localising vaccination services in collaboration with welfare groups raises opportunities for the on-going CVP and other immunisation programmes, and constitutes an opportunity for ethnic and religious minorities to collaborate in safeguarding community health.

## Introduction

Ethnic and religious minorities in England have been disproportionately affected by SARS-CoV-2^1^ and are also widely reported to be less likely to accept vaccines offered as part of pandemic control measures.^2-3^ High incidences of SARS-CoV-2 have been observed among Orthodox Jewish neighbourhoods in the UK,^4^ the US,^5^ and Israel.^6^ This disproportionate burden of SARS-CoV-2 among Orthodox Jewish neighbourhoods should be understood against a backdrop of previous outbreaks of vaccine-preventable disease, especially measles, which have been attributed to suboptimal vaccination coverage rates.^7-10^ In 2018-19, the largest measles epidemics in a quarter century were recorded in the US and Israel, which originated in New York and were linked to unvaccinated Orthodox Jews travelling between the two jurisdictions.^10-12^

The World Health Organization ranked vaccine hesitancy as among the top ten threats to global health in 2019, commensurate with the dangers posed by climate change and antimicrobial resistance. As in any population, concerns of vaccine safety exist in Orthodox Jewish families.^9,13^ There is evidence to suggest that a small but globally networked group of non-vaccination activists are targeting Orthodox Jewish neighbourhoods to propagate vaccine safety concerns.^14-15^ Yet, the contemporary public health and scholarly emphasis on hesitancy is in danger of focusing attention on people as problems, rather than examining the accessibility, efficiency and responsiveness of services in minority settings.

Studies consistently report that Orthodox Jews in the UK and Israel have suboptimal access to vaccination services, often because of practical issues with large family sizes,^16-17^ which indicates structural issues in service delivery and equity. Recognising that a ‘one size fits all’ approach would not improve vaccination coverage rates in Orthodox Jewish neighbourhoods in North London, a WHO Tailoring Immunization Programme (TIP) was conducted in 2014 to diagnose barriers and enablers to vaccination and recommend evidence-informed responses to improve uptake.^16^

The present study focuses on attempts to optimise the UK coronavirus vaccination programme among Haredi Jews, who are often and problematically termed ‘ultra-’ or ‘strictly Orthodox.’ Haredi Jews share a commitment to living within the parameters of Jewish law (*halachah*), but form diverse movements characterised by lineage and nuances in customs, stringencies and rabbinic authority. Haredi Jews are self-protective and carefully manage encounters with broader society,^18^ which may raise implications for healthcare services.^19^ Jewish law does not explicitly endorse or prohibit vaccination, though rabbinic authorities generally accept vaccination as a safe way to protect child and population health.^20^ Jewish law, however, is not always the primary influence on vaccine decision-making among Haredi Jewish parents.^13,21^ It should not be assumed that Orthodox and Haredi Jews will defer family health decisions to religious authorities.^22^

### Conceptual framework

Recent studies exploring uptake of SARS-CoV-2 vaccination among Orthodox and Haredi Jewish neighbourhoods in the US report lower likelihood of accepting vaccination and higher rates of hesitancy, and report that primary disseminators of vaccine information should be trusted stakeholders.^23^ Considering how convenience and confidence affect use of vaccination services in this ethnic and religious minority, we examine the possibility of localising services in settings that are affected by outbreaks of vaccine-preventable disease. Emerging directly from the data analysis in our study, we define localising vaccination services as collaborating with community health organisations to deliver vaccines in ways that meet the situated (context-specific) needs and expectations of minority groups.

Our premise is that localising vaccination services requires a broader conceptualisation of convenience (a recognised influence on vaccine uptake), defined as ‘the degree to which vaccination services are delivered at a time and place and in a cultural context that is convenient and comfortable.’^24^ We explore the potential for vaccination services to not only be tailored in a convenient and culturally appropriate manner, but localised and co-delivered with welfare groups that are valued, trusted and managed within minority settings. We suggest that such an approach might simultaneously promote confidence and remove barriers to accessing routine vaccination services and future coronavirus vaccines.

Localising vaccination entails more than attempts to partner with religious and communal authorities, which are recommended in global health delivery strategies,^25^ especially as part of mass vaccination programmes in the global south such as polio.^26-27^ Rather, it involves upscaling public health relations with welfare services operated by and for minority groups.

To illustrate a localised vaccination collaboration, we examine the case of Haredi Jewish emergency services that partnered with local health authorities to implement initial coronavirus vaccine drives within their neighbourhoods across the UK. As part of this analysis, we explore perceptions of localised vaccination collaborations from the perspective of public health services and the intended beneficiaries.

## Methods

This qualitative research was conducted ancillary to a study examining rates of SARS-CoV-2 seroprevalence in a Haredi Jewish population in the UK.^4^ Methods consisted of semi-structured informal interviews and discursive analysis of Haredi print media and social media pertaining to the UK national coronavirus vaccination programme. A key strand of the interviews focused on the involvement of ‘*Hatzola*’ in the provision of coronavirus vaccines. *Hatzola* (rescue, save) is a volunteer emergency medical service instituted by and for Haredi neighbourhoods around the world. Haredi Jews view Hatzola as a culturally-appropriate service because their staff and volunteers are Haredi and can communicate health information and respond to questions in vernacular languages (English, Yiddish). Hatzola personnel also wear uniforms and *kippot* (religious head covering for males), which distinguish them as members of the Haredi community.^28^ The London Hatzola branch supported the local coronavirus pandemic response by providing emergency care and public health messages, and hence offered a case study to examine the localisation of vaccine services.

We recruited 28 participants from professional networks, past research projects and via snowball sampling. Interviews lasted between 30 and 90 minutes and were recorded with participant consent. The participants were grouped into three key research clusters; i. Public health (PH), 8 professionals based in Clinical Commissioning Groups (CCGs), Local Authorities, the NHS and Public Health England; ii. 10 community welfare and religious representatives (CR); iii. and 10 household members (HM). The first cluster included public health professionals serving the Jewish neighbourhood, and the second comprised local rabbinic authorities, leaders of community welfare organisations, Jewish healthcare professionals, and public relations representatives. Household members ranged in age, gender, educational and professional background, and the Haredi movement to which they affiliate.

The interviews were conducted by BK and TC using online video conference software, or by telephone for a minority of community participants without home access to the Internet, and detailed notes were made. Interviews were conducted between February and May 2021. The interviewers used topic guides that included questions about participant responses to the coronavirus pandemic and their views on the control measures, including the UK’s national coronavirus vaccination programme. This paper presents data from the analysis of vaccine-related data.

Analysis of the data was inductive and thematic, whereby theoretical insights emerge from prolonged engagement with the data rather than being pre-conceived.^29-30^ The data was analysed by BK and TC, who initially coded the same 6 transcripts as a test of reliability. The results were situated in BK’s long-term ethnographic investigation into public health relations with Haredi Jewish minorities^13,19^ and TC’s research examining barriers to accessing vaccination services.^16^ All names of participants, their precise PH roles, and their locations have been anonymised to protect their identities. Ethical approval to conduct this study was provided by the London School of Hygiene & Tropical Medicine (reference: 22532).

## Results

Our attention to localised vaccination services raise five key considerations: i) pre-pandemic collaborations to address issues in routine vaccination delivery; ii) scaling up collaborations during the coronavirus vaccination programme and division of responsibility; iii) benefits and limitations of a localised approach for minorities with historically suboptimal levels of vaccination coverage; iv) household responses to Hatzola-hosted coronavirus vaccination sessions; and v) coronavirus vaccine information and responding to situated concerns.

### i. Pre-pandemic collaborations to address issues in routine vaccination delivery

Hatzola’s role in the coronavirus vaccination programme evolved from their involvement in past PH campaigns to promote routine childhood vaccination. Historically suboptimal levels of childhood vaccination meant that PH workers recognised the value of sustaining relations with Jewish representative bodies, rabbinic authorities, and local healthcare professionals. Established relationships were drawn upon when major outbreaks of measles emerged in Haredi communities in London following the 2018-19 epidemics. PH organisations introduced outbreak response meetings, and Hatzola was commissioned to produce information about vaccine uptake for publication in Haredi newspapers and circulars. Hatzola was considered better placed to deliver targeted – often direct – messaging about the dangers of measles infection, and to quote one consultant, ‘*the sort of comms that we can’t necessarily put out as a public health team*’ (PH1). Commissioning Hatzola for a support role during the measles outbreaks then provided the basis to upscale their involvement in the coronavirus vaccination programme.

PH professionals confirmed that the findings from WHO TIP study (running between the years 2015-16)^16^ reflected the issues they encountered in seeking to increase vaccine uptake. They stated that access and convenience (e.g. need for flexible clinic times to cater for large families) continued to be the primary issues impeding vaccine uptake rather than vaccine hesitancy. They also added that there was ‘*a lack of health knowledge in the community*’ (PH1), which needed to be addressed to promote the value of vaccination in protecting child health. Against the backdrop of the 2018-19 measles outbreaks, some public health workers suggested that improving vaccination coverage rates required greater input from Haredi Jews themselves:

‘*I think it’s better if it’s delivered for them [community] by them maybe. Or at least with them*’ (PH2)

Yet, maintaining sustainable sources of PH funding to continue vaccination programmes in ways that met the expectations of Haredi parents was described as a long-running problem. A clinician noted how staff dedicated to ‘call and recall,’ who could monitor cases of non-vaccination, such as not attending scheduled appointments, were discontinued, as was the involvement of health visitors in supporting routine vaccination programmes. Inconsistent service provision due to funding limitations was considered by PH staff to engender mistrust on the part of parents making vaccination decisions. PH staff recognised the need for additional financial resources from the central government to deliver vaccinations in minority settings:

‘*As we say for the 10% of the population or 15% of the population that won’t get vaccinated, you have to think differently and it will cost you more money*’ (PH2).

### ii. Scaling up collaborations with Hatzola during the coronavirus vaccination programme and division of responsibility

The scaling up of Hatzola’s involvement in the coronavirus vaccine programme was made possible by their role in promoting routine immunisations, and was a direct by-product of providing emergency care and circulating public health guidance to Haredi neighbourhoods throughout the pandemic. It was agreed that a select number of Hatzola-led vaccination sessions would take place in one of the designated and approved local vaccination centres, with a clear division of roles between Hatzolah and PH bodies.

Hatzola hosted these vaccination sessions and had responsibility for promotion, distributing appointments to callers and administering vaccines. Events were also supervised by Jewish healthcare professionals working in the community, which offered continuity between delivery of routine vaccinations and the coronavirus vaccine programme. One healthcare professional suggested that this collaboration meant that Haredi Jews were not just intended beneficiaries of a national vaccination programme, but also had a stake in how the programme was being delivered:

‘*Public health sort of handed over - at least sufficiently - the front of the campaign to Hatzola so it [*…*] was actually coming from the Haredi community rather than being imposed on it*.’ (CR8)

PH staff stated that they managed the sessions from ‘behind the scenes’ (PH3), which meant that PH teams in the UK, maintained responsibility for logistics, training vaccinators, and accurate vaccination records. PH professionals trained 8 male Hatzola volunteers to administer vaccines (and 2 female Jewish healthcare assistants) and ensured sessions operated in accordance with COVID-19 compliance. Physical distancing was, however, reported to be a difficult issue to control in a tightly-knit neighbourhood setting.

Scheduling appointments was an example of how both parties collaborated with designated roles. HMs were requested to phone Hatzola to make appointments in advance (which was beneficial to people without internet access at home), and records were maintained by CCGs:

*‘*…*we [PH professionals] needed to control the appointment book so that we could keep track of who actually got jabbed, they [administrators] would then pass that information to us and we would create our own appointment book internal to the system*.*’* (PH3)

PH professionals encountered some challenges in working in an ethnic and religious minority, which the collaboration with Hatzola helped to address. One challenge involved retrieving names with variations in spelling, and interpreting dates of birth across the Gregorian and Hebrew calendars.

All Hatzola responders are male, which raised questions among PH professionals about the gender of vaccinators for a religiously conservative minority, characterised by a strict separation of men and women on the basis of modesty. While provisions for female vaccinators were made in the Hatzola-hosted events, this turned out not to be an issue since only a minority of attendees requested this service:

*‘So, we had a male/female choice for people if they wanted it, although interestingly I think probably only two people wanted to be vaccinated by a person of the same sex. Most people were not bothered. I thought there would be a greater requirement*.*’* (PH3)

Hatzola-hosted vaccination events in other UK regions arranged separate sessions for men and women to make services user-friendly. Hence in other regions, the gender of the vaccinator was seen as less important than the mixing of male and female attendees when attending sessions. Adopting this localised and collaborative approach indicated how the issues perceived by PH professionals to influence vaccination service delivery are not always a major problem in practice.

### iii. Opportunities and challenges of a localised approach for minorities with historically suboptimal vaccination coverage

The involvement of Hatzola in local vaccination delivery was perceived by CRs and PH professionals as increasing trust in the national coronavirus vaccination programme.

‘*Knowing that Hatzola is sort of sponsoring this. Knowing that you’re going to a place that is going to be – have people around you who are like you, who you trust. Having them vaccinate you. It seems to make all the difference*.’ (PH3)

A CR professional who lived in the neighbourhood and was involved in raising awareness of the Hatzola-hosted vaccination sessions expanded further on the issue of trust. They claimed that Hatzola’s branding helped convince those who would otherwise not have received or who may have been hesitant to accept the vaccine:

*‘Now I’m sure there might be a minority of people that wouldn’t have taken the vaccine and have taken it now. people saw Hatzola’s logo, or Hatzola brand and Hatzola events, that in itself was a strong enough message for people to trust they can come and they can take the vaccine and that it’s safe*.*’* (CR7)

Closely aligned to trust, was the perception that Hatzola are viewed as an acceptable and celebrated service across a diverse minority. Rabbinic authorities felt this was partly because Hatzola volunteers themselves represent different branches of Haredi Judaism:

‘*Hatzola I think is accepted across the board of the Haredi community. They themselves, their members are drawn from the full spectrum and I think they are seen as heroes*.’ (CR2)

The demand for the number of appointments met availability, indicating the popularity of this localised approach to vaccination delivery, as one public health worker noted:

‘*We had a queue down the road and around the corner over and above all the people we’d booked in*.’ (PH3)

PH professionals were, however, keen to assert that convenience was not only an expectation of Haredi Jewish constituents, and constituted a challenge of the UK coronavirus vaccination programme:

*‘Our feedback from every community, not just the Orthodox community, is “I don’t want to go trekking off to ExCeL, having to get on the bus or on the tube,” so can’t you come to me?’* (PH3)

PH professionals did, however, recognise that localised vaccination services offered particular benefits to encourage uptake among Haredi Jews. Haredi Jews were viewed by a PH professional as a self-protective minority that felt uncomfortable ‘integrating with others,’ so Hatzola-hosted vaccination services were perceived as an ‘Orthodox-friendly place to come’ (PH3).

Participants were careful to note that Hatzola’s involvement was not intended to replace the role of mainstream coronavirus vaccination sites, but rather to initiate awareness of – and ‘normalise’ (HM4) – the vaccine programme:

*‘And it [the pandemic situation] probably would have fed off that people would have had the vaccination even without Hatzola, but the fact that there is – it generates a fervour and an excitement in the community, “oh, Hatzola has managed to get it at this,” you know, they have a higher authority. All that gives a sense of we’re part of it*.*’* (HM3)

This position is reflected in the select numbers of Hatzola-hosted sessions that have been organised during the UK coronavirus vaccination programme:

‘*Even whilst Hatzola vaccination events were happening, people were still getting their invitations in the post from NHS and just going wherever they were told to go*’ (CR7).

The majority of Haredi Jewish constituents were then expected to attend general coronavirus vaccination sites for the first and/or second dose of the vaccine (with Hatzola maintaining a role in allocating vaccination appointments to general sites). As one local healthcare professional responded:

*‘It was a really good move and really encouraged the uptake of vaccines, not just for those particular events, I think we held four events, but that actually got the community into the idea that this is something we want to do*.*’* (CR8)

Localised vaccination services also involved flexibility when it came to who was eligible for vaccination, which, at the time these events took place (February 2021), should have been older people, clinically extremely vulnerable groups, carers and workers in public facing charitable or voluntary roles. As a healthcare professional noted:

‘*It should be kept in mind though that this was partly successful because the rules as to who could get the vaccine were interpreted a little bit more loosely at the time*.’ (CR8)

While deviating from national protocols in place due to limited supplies of coronavirus vaccines, this approach provided flexibility – especially if slots had remained unfilled or cancelled – and meant that localised vaccination services were able to boost community coverage rates. Several healthcare professionals were keen to learn from this collaborative effort with the aim of improving uptake of routine vaccinations:

‘*I would like to see one of the outcomes of this epidemic is the – just the strengthening of the childhood immunisations*.’ (CR8)

### iv. Household responses to Hatzola-hosted coronavirus vaccination sessions

Hatzola was not the only influence on coronavirus vaccine decision-making. Household members valued the opportunity to protect themselves, and to plan flights to visit family members amidst public health discussions of requiring a ‘vaccine pass’ for travel. However, the trust invested in Hatzola by Haredi Jewish constituents signals the benefits of a localised approach for minorities with historically suboptimal of vaccination coverage:

*‘If they see Hatzola doing it they’re much more likely to go and have it done. It’s the way the community is built in terms of trust. The Jewish community is very knowledgeable in terms of, that they will ask a lot of questions. They don’t just take things at face value, yeah. So, just because Public Health England comes out and says you should have a vaccine, they will not just go and vaccinate. They need a lot of information before they put trust in something*.*’* (HM5)

However, the Hatzola-hosted events were not universally considered to invest trust in the coronavirus vaccine especially among household members who refused childhood vaccinations:

*‘Hatzola’s already done a vaccine drive here, it doesn’t impress me. Until I know what the side-effects are, I’m not interested. They’re claiming to find the vaccine very quickly but we don’t know the side-effects*.*’* (HM8)

Household respondents who were concerned about the safety of the coronavirus vaccine criticised Hatzola for collaborating with the vaccination programme, but were cautious about the pandemic and careful to maintain physical distancing:

*‘Hatzola’s actually telling them, “Go get vaccinated. Go get vaccinated*.*” For me, everything is like completely the opposite. They’re not being careful. I would be careful with social distancing, but I just think that I’m afraid of this vaccine. We’ve got all the vaccines until now, all the boosters. This one feels wrong to me, so I’m going to keep away as long as I can*.*’* (HM6)

The positions of these parents reflect how, in all populations, there will be a small proportion of people opposed to vaccination – which does not undermine the potential value of localised services for the wider population.

### v) Coronavirus vaccine programme information and responding to situated concerns

A wide range of stakeholders from the PH and CR research clusters were involved in promoting local coronavirus vaccination uptake through information and endorsement initiatives. Public relations campaigns decided to amplify the authority of Hatzola as a vehicle to mediate public trust:

*‘We decided not to engage in trying to convince people that conspiracies are not true. The very fact that we decided to put out adverts telling people to get vaccinated, that people saw Hatzola’s logo, or Hatzola brand and Hatzola events, that in itself was a strong enough message for people to trust they can come and they can take the vaccine and that it’s safe*.*’* (CR7)

Rabbinic and communal authorities implicitly and explicitly endorsed the Hatzola-hosted sessions and general coronavirus vaccination programme. It should be noted that to our knowledge no official mandate or ruling of Jewish law (*psak halacha*) was put forward by Haredi rabbinic authorities in the region under study, to the concern of some healthcare professionals involved in delivering the coronavirus vaccines:

*‘Rabbis felt it was a health matter and it was between the individuals and their own doctors to make the decisions. They did not feel that that was a halachic issue, that it was a medical issue within which they shouldn’t interfere*.*’* (CR8)

Rather, rabbinic authorities took an approach of discussing the vaccine on a one-to-one basis with constituents, though it is important to note that senior Haredi rabbinic authorities were photographed being vaccinated as part of the PR work around the Hatzola events. For participants affiliated to particular Haredi movements with rabbinic leaders based in the USA or Israel, health decision-making was influenced by the guidance of local and international rabbis.

Situated concerns around coronavirus vaccines did arise at a household level, which PH and CR sought to address through local information campaigns. HMs commented on the circuitous flow and exchange of vaccine safety information from the US and Israel via WhatsApp groups. Unsubstantiated claims that coronavirus vaccines could affect women’s fertility were cited by HMs, which raised particular implications in a setting where larger family sizes are idealised:

*‘There are rabbonim in America not letting the girls do vaccines due to fertility issues. So I don’t know, at the moment, I wouldn’t, due to fertility issues, give her [daughter] a vaccine yet, no, I definitely wouldn’t*.*’* (HM8)

Other women were concerned by claims that coronavirus vaccinations could affect fertility, but ultimately ‘*put faith in the fact that it was safe*’ (HM10). PH workers had encountered concerns that the vaccines could affect fertility and were working to reassure younger age cohorts through targeted communications campaigns. Moreover, a challenge for public health collaborations was directly addressing the issue of infertility, but within the confines of modesty standards:

‘…*how do we get the information out there in a culturally sensitive way, in public literature, that’s not going to offend people, we don’t want those kind of subjects spoken about*.’ (HM5)

Media reports of extremely high seroprevalence of SARS-CoV-2 among Haredi Jews in the UK^4,31^ meant that healthcare professionals had to assert that vaccination was still important even if HMs had previously been infected:

*‘We were asked time and time again, “if the prevalence rate was so high, aren’t we all immune, don’t we have herd immunity?” We’ve had to spend quite a lot of time explaining why that isn’t the case*.*’* (CR8)

Hence, promotion of a national vaccination campaign had to consider the situated issues that arose at local-levels.

## Discussion

Implementing a localised model of vaccination delivery enabled public health services to engage a self-protective religious minority through a trusted care provider, which took an active role in communication, booking and hosting vaccination sessions in a designated clinic. Hatzola served as a suitable partner as it is operated by and for Haredi Jewish minorities and holds their trust as a provider of rapid response services. Moreover, Hatzola was already involved in the local epidemic response by providing emergency care and public health messages. Localising vaccination services required flexibility in implementation to promote higher local coverage rates, which reflected practices observed in the national coronavirus vaccine programme – such as re-distributing doses to avoid waste.^32^

The localised approach involved hosting a select number of Hatzola-led vaccination sessions to serve as a gateway for household members to be exposed to the national coronavirus vaccination programme in a space and service that was perceived as familiar, trusted and convenient. Subsequently, household members could book vaccination appointments at local vaccination centres *through* Hatzola. The region under study is home to a diverse range of ethnic minorities, and like many London Boroughs, coronavirus vaccination coverage is lower than the national average.^33^ The continued collaboration between public health professionals and Hatzola offers an opportunity for decision-makers to identify whether uptake remains suboptimal among Haredi Jews, whether additional Hatzola-hosted sessions are required, and how these could support the next phase of the national coronavirus vaccination programme.

Unlike past studies,^16^ our data suggests that the coronavirus vaccination was valued for international travel. Yet, a minority of participants maintained, questions and concerns around vaccination safety, which were not addressed by the Hatzola-hosted sessions. The issue of refusal in a minority of participants likely reflected similar issues in achieving universal coronavirus vaccination coverage across the UK population,^2^ which includes public concerns around safety.^34^ While we did encounter misinformation about vaccinations in general, it is important to remember that coronavirus vaccines were developed and implemented at record-pace. Hence, it is not surprising that some household members cited concerns about the safety and long-term effects of new coronavirus vaccines.

Policy-makers have long considered public health collaboration with religious and communal authorities an important part of effective and sensitive vaccine delivery-strategies.^25^ Moreover, the WHO TIP programme is premised on community engagement, which ‘means including members of underserved population groups among active stakeholders who will define barriers to immunization and design solutions to overcome them.’^35^ The UK coronavirus vaccine programme has since heralded innovative attempts to make vaccines more accessible for minority groups, evidenced by administering vaccines in ‘pop-up’ clinics in places of worship and community centres.^36^ The model of localising vaccination that we outline builds on previous learning around issues of vaccine confidence, convenience and complacency by sharing responsibility for vaccine delivery with trusted community services.

### Localising vaccination services: investment, engagement and coordination

Based on our analysis of a localised delivery of the UK coronavirus vaccination programme, we suggest there is a space to discuss complementary vaccination services to improve access to routine vaccinations among ethnic and religious minorities with a history of suboptimal coverage leading to outbreaks of preventable disease. A model of localised vaccination services does not mean overhauling vaccination services entirely, but identifying how gaps in service provision can be addressed in ways that involve trusted community organisations.

Localising vaccination services in ways that enable collaboration and coordinated delivery is a promising approach, which is not cost neutral. Challenges arise when funding is not allocated for localised services, but sustained investment may outweigh costs in minority settings that are susceptible to outbreaks of preventable disease. In the model of localised services that we discuss, a degree of responsibility over vaccine location, delivery and promotion is handed to partnering community organisations. Yet, collaboration with minority welfare groups requires considerable public health oversight. Our analysis suggests that public health services, local authorities and central government will need to maintain responsibility for assessing the suitability of partnering organisations to maintain public trust in vaccination, legalities of administration^37^ and accurate data recording, procurement of vaccinations, purchasing, cold storage, oversight/accountability of collaborative groups, signposting sites for reporting suspected side effects for reporting suspected side effects (e.g. United Kingdom: Yellow Card: USA: Vaccine Adverse Event Reporting System).

Future localisation of vaccination services could take two key approaches of collaboration via communication or implementation strategies. A communications-only focus could promote state vaccination programmes in ways that directly address the concerns of ethnic and religious minorities, as Hatzola did in the measles outbreaks described above. An implementation method is a more complicated operation, indicated by the scaled-up involvement of Hatzola in the UK coronavirus vaccine programme, and involves the following key considerations:

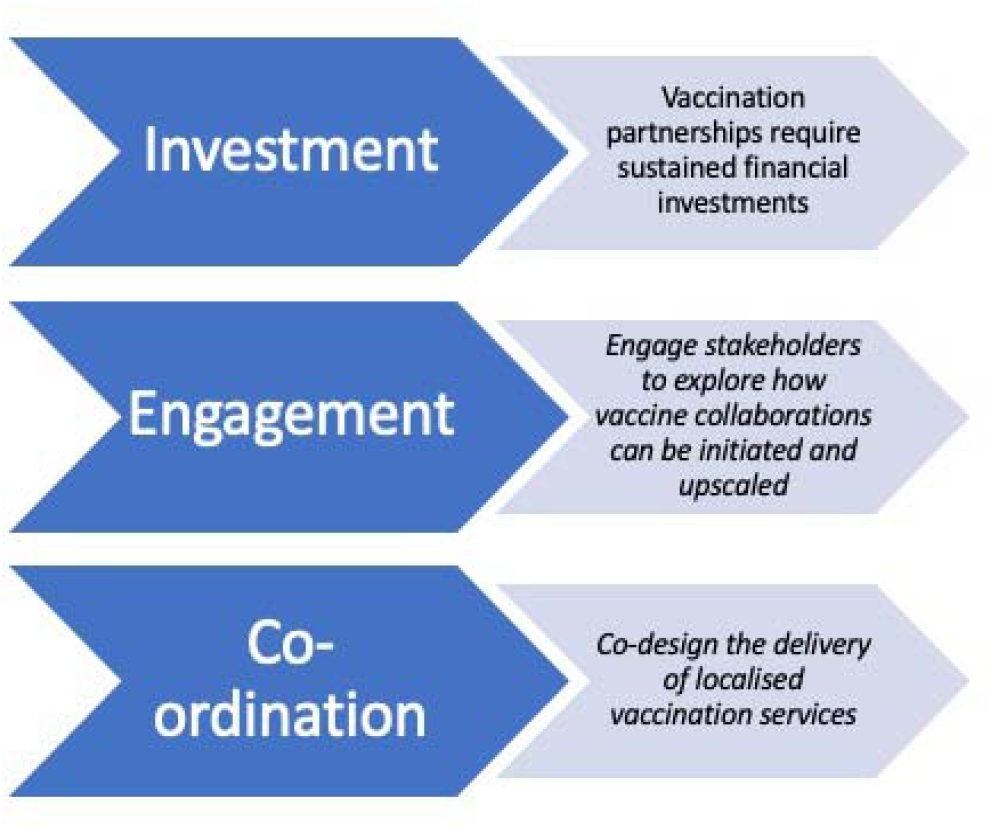

### Strengths and limitations

UK data has consistently suggested that ethnic and religious minorities are less likely to accept the new COVID-19 vaccinations.^2^ This study interviewed a wide range of people, including public health professionals, community representatives and intended beneficiaries to examine opportunities to promote high coverage levels.

We recognise that some stakeholders involved in delivering the coronavirus vaccination programme were unable to be recruited. Further work should consider how collaborative organisations perceive the feasibility of localised vaccination services as outlined above.

## Conclusion

This study examined how a national vaccination campaign, the largest in British history, was localised in collaboration with welfare groups, which raises implications for subsequent coronavirus booster shots as well as the routine vaccination programmes. Localising vaccination services raises opportunities for greater vaccine equity by supporting ethnic and religious minorities to collaborate in safeguarding community health and wellbeing.

## Data Availability

The qualitative data collected is sensitive and we did not obtain ethical approval to share this data beyond the research team.

## Author contributions

TC, MM and BK conceived of the study. BK and TC planned and conducted the qualitative data collection and led the data analysis. KG, ChR, RE, NS, LL, SMJ contributed to the design of the study. All authors reviewed the analysis and contributed to writing the manuscript.

## Ethics

Ethical approval to conduct this study was granted by The London School of Hygiene and Tropical Medicine Research Ethics Committee (Ref: 22532).

## Acknowledgements

We thank our study participants for their time and insights, and Dr William Waites for comments on an earlier draft.

